# Utilisation of Remote Capillary Blood Testing in an Outpatient Clinic Setting to improve shared decision making and patient and clinician experience: a validation and pilot study

**DOI:** 10.1101/2020.08.03.20167106

**Authors:** Lisa Nwankwo, Kate McLaren, Jackie Donovan, Melody Zhifang Ni, Alberto Vidal-Diaz, Michael Loebinger, Anand Shah

**Affiliations:** Pharmacy department, Royal Brompton and Harefield NHS Foundation Trust, UK; Department of Respiratory Medicine, Royal Brompton and Harefield NHS Foundation Trust, UK; Department of Pathology, Royal Brompton and Harefield NHS Foundation Trust, London, UK; National Heart and Lung Institute, Imperial College London, London, UK; Department of Infectious Disease Epidemiology, School of Public Health, Imperial College London, UK; Faculty of Medicine, Department of Surgery and Cancer, Imperial College London, London, UK

**Author notes:** Joint first authors. Both LN and KM contributed equally to the work.

## Abstract

**Background:** In a tertiary respiratory centre, large cohorts of patients are managed in an outpatient setting and require blood tests to monitor disease activity and organ toxicity. This requires either visits to tertiary centres for phlebotomy and physician review or utilisation of primary care services.

**Objectives:** This study aims to validate remote capillary blood testing in an outpatient setting and analyse impact on clinical pathways.

**Methods:** A single-centre prospective cross-sectional validation and parallel observational study was performed. Remote finger prick capillary blood testing was validated compared to local standard venesection using comparative statistical analysis: paired t-test, correlation and Bland-Altman. Capillary was considered interchangeable with venous samples if all 3 criteria were met: non-significant paired t-test (i.e. p>0.05), Pearson’s correlation coefficient (*r*) >0.8 and 95% of tests within 10% difference through Bland-Altman (Limits of agreement). In parallel, current clinical pathways including phlebotomy practice was analysed over 4 weeks to review test predictability. A subsequent pilot cohort study analysed potential impact of remote capillary blood sampling on shared decision making and outpatient clinical pathways.

**Results:** 117 paired capillary and venous blood samples were prospectively analysed. Interchangeability with venous blood was seen with HbA1c (%), total protein and CRP. Further tests, although not interchangeable, are likely useful to enable longitudinal remote monitoring (e.g. liver function, total IgE, and vitamin D). 65% of outpatient clinic blood tests were predictable with 16% of patients requiring further contact due to actions required. Pilot implementation of remote capillary sampling showed patient and clinician-reported improvement in shared decision-making given contemporaneous blood test results.

**Conclusions:** Remote capillary blood sampling can be used accurately for specific tests to monitor chronic disease, and when incorporated into an outpatient clinical pathway can improve shared decision making and patient experience. Further research is required to determine health-economic impact and applicability within telemedicine-based outpatient care.

## INTRODUCTION

Given the current COVID-19 pandemic situation, there has been a drive to enable remote consultation including virtual internet-based telecommunication and remote monitoring tools.^1^ Tertiary respiratory centres manage large cohorts of highly COVID-19 susceptible individuals from across the whole of the country. With centrally commissioned care and prescriptions of disease-modifying therapy for patients with Cystic Fibrosis (CF), severe asthma or interstitial lung disease, patients often require outpatient monitoring of inflammatory or biochemical markers to determine response/toxicity, alongside therapeutic drug monitoring for antimicrobials. Even prior to the current pandemic, as prognosis has improved, these individuals are often fully independent and lead busy lives in full-time employment making regular hospital-based appointments difficult with added socio-economic cost.^2^

At present, however, there are no suitable easily accessible local facilities to enable remote blood monitoring for NHS providers, and patients often rely on an increasingly stretched primary care resource for routine phlebotomy and monitoring, with resultant adverse socioeconomic impact. In the current COVID-19 pandemic, this is however problematic for individuals with high-risk diseases that require shielding. Even prior to the current scenario, primary care physicians are often uncomfortable monitoring blood tests for patients using specialist drugs and patients often feel helpless in establishing who can and should take responsibility for monitoring. This increases failure demand^3^ (avoidable rework) where patients and/or primary care providers contact tertiary care providers for additional demands adding further strain on NHS resources and delay. In advanced disease, patients with chronic respiratory disease are also often limited by breathlessness and rely on relatives, friends and the NHS transport system to assist them with transportation, particularly when requiring supplemental oxygen. As chronic disease is associated with progressive social isolation, such support networks are often lacking, and likely not available with current shielding guidelines.^4-8^ It is therefore a priority to enable practical blood test monitoring solutions which are not compromised by COVID-19 shielding requirements, social isolation or lack of suitable local facilities.

Within face to face interactions or teleconference consultations, remote capillary blood testing could provide contemporaneous clinical information to aid shared decision making between the patient and healthcare professional and enable efficient clinical review alongside prescription modification. Greater patient involvement has been shown to improve health outcomes and treatment adherence alongside reducing long-term healthcare cost.^9^ Thus, the ability to monitor remotely through capillary blood testing has the potential to streamline outpatient care, ensure better patient care, improve transparency and reduce health-economic burden for patients, primary and tertiary care providers.

In this multi-phase prospective cross-sectional single-centre pilot study, we aim to 1) Evaluate accuracy and usability of remote capillary blood testing in adults with chronic respiratory disease when compared with standard venesection; 2) Analyse the impact on clinical pathways in the outpatient tertiary respiratory setting to reduce failure demand and improve shared decision making.

## METHODS

This was a pilot study aimed at improving shared decision making by facilitating contemporaneous results at the time of outpatient clinic attendance. We adopted applicable elements of the extension of CONSORT 2010 checklist for pilot trials ^10^ for the implementation phase of this study.

### Setting

The setting was a large tertiary referral respiratory centre in London.

### Study design

A parallel phase prospective cross-sectional and observational study was designed.

1. Cross-sectional validation study of finger-prick remote capillary blood testing compared to gold standard (venous phlebotomy in outpatient setting).
2. Analysis of potential clinical pathway impact in outpatient tertiary respiratory care of remote capillary blood testing to reduce failure demand and improve shared decision making.

### Patients and study period

Study subjects were recruited from within the hospital between January 2018 to October 2019. Study materials and protocols were approved by the North West - Haydock Research Ethics Committee (REC reference: 18/NW/0491) with informed written consent obtained at participant’s clinic visit or inpatient stay. 124 patients were recruited over the study period (Table 1). We identified specific patients predicted to require blood tests during their routine clinic appointments. Inclusion criteria were as follows: patients > 16 years of age with chronic lung disease treated at the tertiary respiratory centre, and ability to provide informed consent. Exclusion criteria were: inability to provide informed consent, adults with needle phobia, currently on therapeutic dose anticoagulation or direct acting oral anticoagulants, and abnormal clotting profile or thrombocytopenia (Platelet count < 100×109/ L).

**Table 1:**
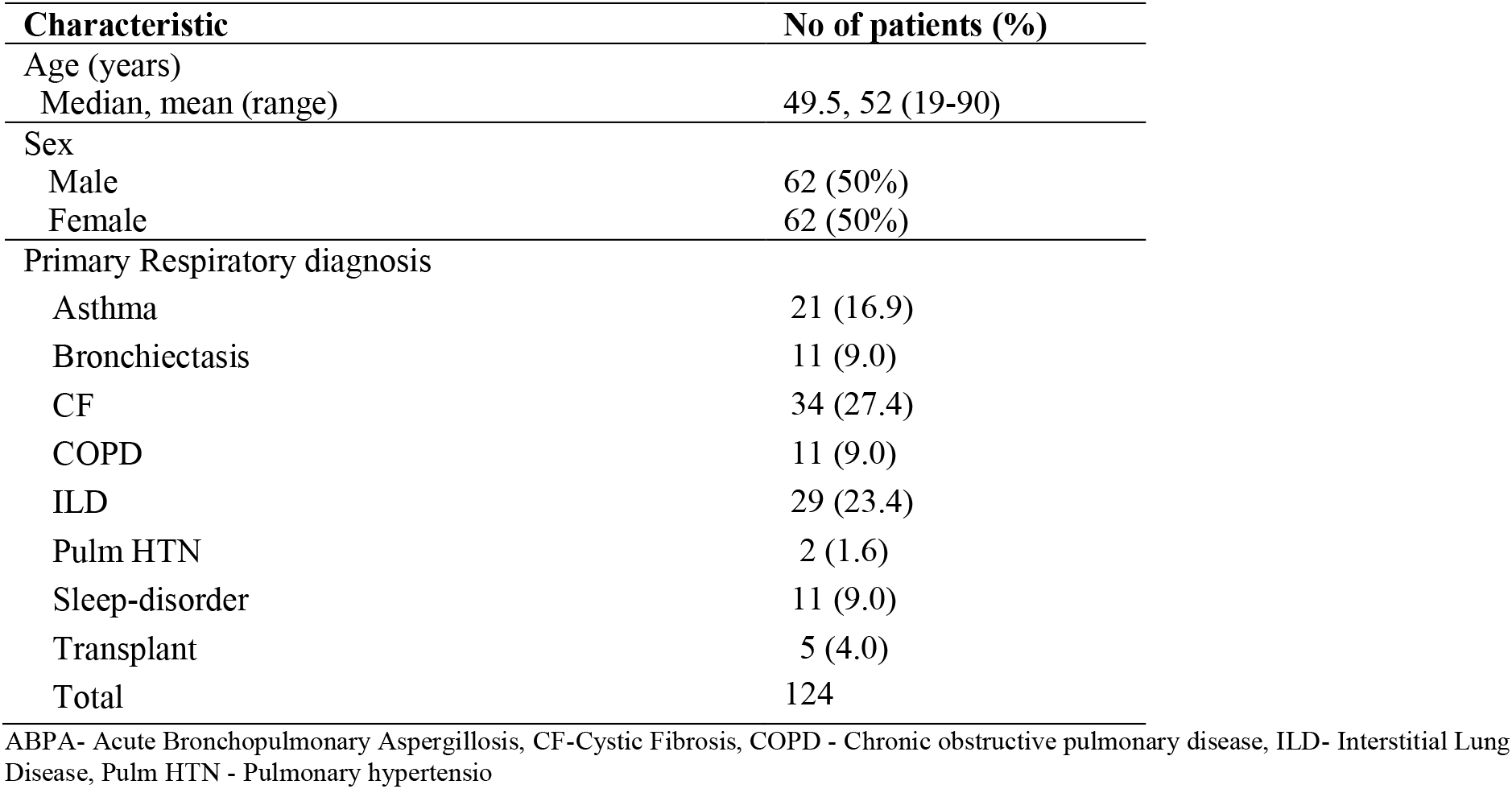
Demographics of the patients involved in the validation study

### Intervention

The study consisted of 2 phases:

### Phase 1

Once recruited, study participants had capillary blood testing performed using a blood sampling kit (*Thriva®)* with CE marked finger-prick (capillary) sampling components by a clinical research coordinator and simultaneously by venesection. To imitate remote testing, capillary samples were posted (using standard first-class postage and custom UN3373 compliant transport packaging). Outpatient phlebotomy samples were sent to the laboratory as per standard clinical care. All samples (capillary and venous) were analysed at the hospital laboratories. Haemolysis index value and time in postal transit data was collected. As usually only one 0.5ml or 0.6ml tube can be filled through capillary blood sampling, two groups of tests were determined based on clinical requirement as follows:

Group 1: Urea and Electrolytes, Liver function tests, C-reactive protein (CRP), Total Immunoglobulin E (IgE), Vitamin D
Group 2: Full Blood Count, HbA1c

Following capillary blood testing and venous phlebotomy, study participants completed a Net Promoter Score questionnaire^11-13^ evaluating usability of the finger-prick capillary sampling device.

### Phase 2

Simultaneously to the validation study, a prospective analysis of 4 weeks of specialist respiratory clinics was undertaken to identify indication and use of blood tests in an outpatient clinic setting. A Clinical pathway mapping and clinical decision tree were formulated to evaluate impact of blood results on outcome and clinical management.

### Implementation

Following Phase 1 analysis, distinct patient groups were identified who might benefit from pre-emptive capillary blood testing. These patients were approached by phone and enrolled in a pilot study to incorporate remote capillary blood testing into their routine clinic review. Over a 2 week period study subjects were instructed to perform capillary blood sampling prior to clinic review at home, and mail it to the hospital laboratories where the results would be processed in preparation for review. Study subjects and clinicians provided written feedback on the impact of contemporaneous blood results on shared decision making and personal experience in the outpatient setting.

### Statistical analysis

Results were presented as mean ± standard deviation, and median for numerical tests. Tests for normality were applied to the pairs of variables, and non-parametric statistics were applied to those with non-Gaussian distribution. To determine agreement between the blood collection methods a combination of 3 tests were performed: 2-tailed paired t-test, correlation analysis (strength of association) and Bland-Altman analyses (measurement of agreement).^14 15^ For the paired t-test, P values <0.05 were deemed statistically significant meaning they show there was a difference between the two measurements.

Bland-Altman analysis measures the degree of agreement between 2 methods but does not say if these differences are acceptable or not. We have thus set clinically acceptable limits as 10% difference between the two measurement, with *a priori* requirement of 95% of results having to be within 10% difference.

An algorithm was developed to determine the clinical usability and interchangeability of the 2 methods of blood sampling (capillary and venous) (Figure 1), with 1 point scored for each statistical test. We considered a non-significant paired t-test analysis (i.e. p>0.05), Pearson’s correlation coefficient (r) greater than 0.8 and 95% of measurements within 10% difference between capillary and venous test results on Bland-Altman analysis to indicate interchangeability between the two methods.

**Figure 1:**
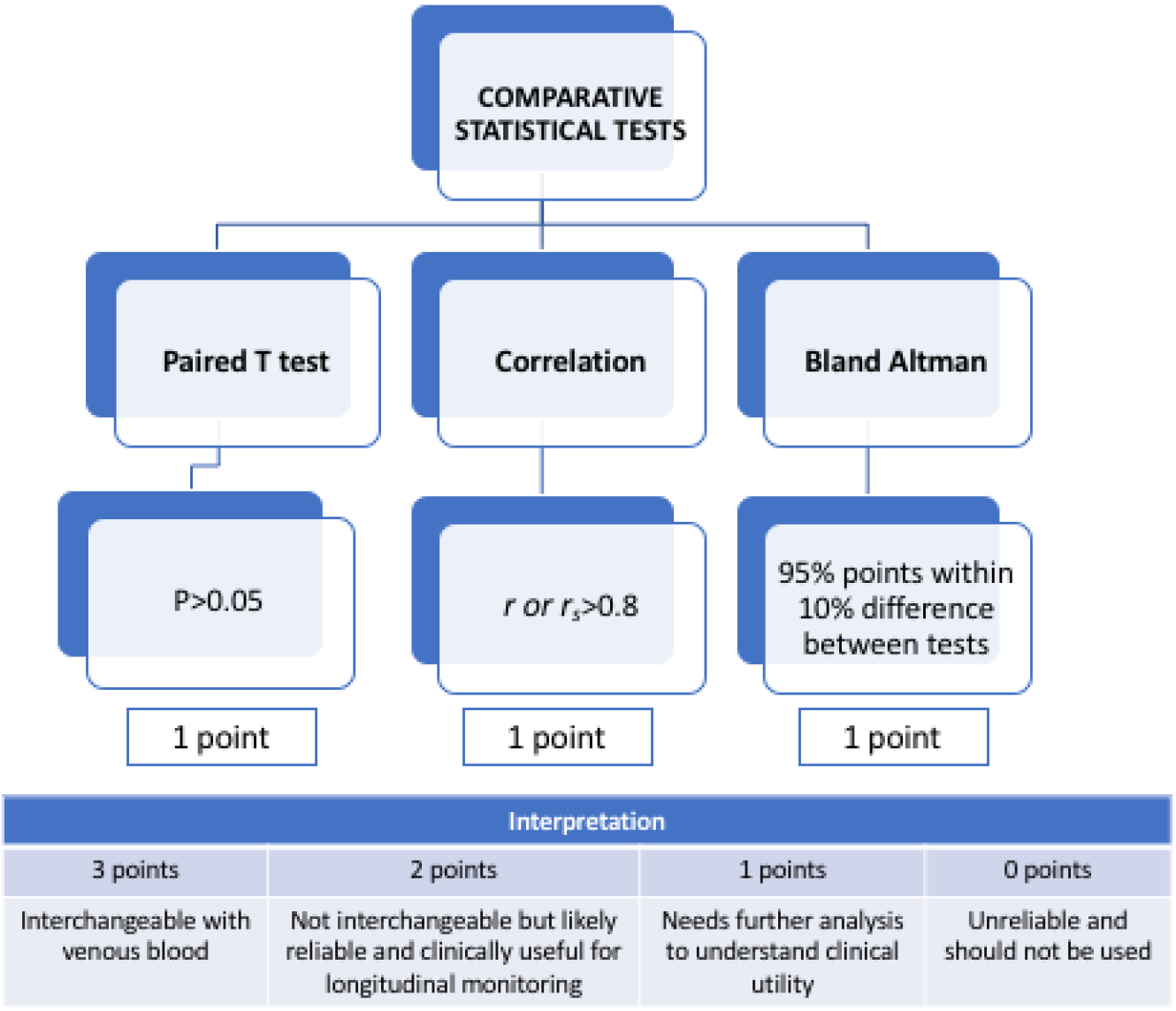
Algorithm for determination of clinical acceptability of capillary tests. 3 points indicates venous blood is interchangeable with capillary blood in the clinical setting, 2 points indicates usefulness for longitudinal monitoring, 1 point indicates care is needed for interpretation of results, and 0 points indicate that the capillary test is unsuitable for clinical use. ‘Bias’ is the average of the differences between the two methods of blood sampling, expressed as a percentage %. Foot note: P= P value. P<0.05 indicates there is a statistically significant difference between the two measurements *r*=Pearson’s correlation coefficient. r_s_ =Spearman’s Rank Correlation Coefficient

To determine whether postal transit time or capillary sample Haemolysis index value had an impact in causing significant differences between capillary and venous sample readings, a Coefficient of variation (CV) analysis, expressed as a %, was applied to some tests that had a low interchangeability score of 1 (See Figure 1 and Table 2 for point scoring explained). The CV is a measure of the dispersion or spread of data points around the mean and is useful in comparing the degree of variation from one data series to another; in this case capillary and venous readings.

**Table 2:** Analysis of remote capillary versus venous phlebotomy in study subjects

| Test | N° of paired samples | Paired T test |  |  |  |  | Correlation |  | Bland Altman |  |  |  | Reference range of test | Interchangeably in clinical setting based Paired T test p>0.05 | Interchangeably in clinical setting based on *r* > 0.8 | Interchangeably in clinical setting based on small Bias OR max 10% difference between tests | Point scoring for clinical acceptability |
| --- | --- | --- | --- | --- | --- | --- | --- | --- | --- | --- | --- | --- | --- | --- | --- | --- | --- |
|  |  | Mean of difference | SD | SEM | 95% CI | Two-tailed P value | *r* (or *r*<sub>s</sub>) | 95% CI | Bias (%) | SD of bias (%) | 95% Limits of Agreement (%) |  |  |  |  |  |  |
|  |  |  |  |  |  |  |  |  |  |  | From | To |  |  |  |  |  |
| Hb | 30 | -0.20 | 11.49 | 2.10 | -4.49 to 4.09 | 0.9247 | 0.73 | 0.50 to 0.86 | 0.12 | 8.15 | -15.9 | 16.1 | 115 - 151 g/L | Yes | No | No | 1 |
| WCC | 29 | -0.69 | 1.16 | 0.22 | -1.14 to -0.25 | 0.0033 | 0.92 | 0.83 to 0.96 | 7.88 | 12.99 | -17.59 | 22.34 | 5.1 - 11.4 10<sup>9</sup>/L | No | Yes | No | 1 |
| Platelets | 30 | 38.67 | 39.80 | 7.27 | 23.80 to 53.53 | <0.0001 | 0.93 | 0.86 to 0.97 | -15.79 | 16.96 | -49.03 | 17.44 | 147 - 397 10<sup>9</sup>/L | No | Yes | No | 1 |
| HbA1c (%) | 31 | -0.05 | 0.20 | 0.03 | -0.12 to 0.026 | 0.2024 | 0.98 | 0.97 to 0.99 | -0.80 | 3.43 | -5.92 | 7.53 | 4.0 - 6.0% | Yes | Yes | Yes | 3 |
| HbA1c (IFCC) | 32 | -0.47 | 2.05 | 0.36 | -1.20 to 0.27 | 0.2024 | 0.99 | 0.97 to 0.99 | 1.21 | 5.44 | -9.46 | 11.88 | 20 – 42 mmol/mol | Yes | Yes | No | 2 |
| Sodium | 36 | -2.22 | 2.57 | 0.43 | -3.09 to -1.35 | <0.0001 | 0.80 | 0.64 to 0.90 | 1.57 | 1.81 | -1.98 | 5.13 | 133 – 146 mmol/L | No | No | Yes | 2 |
| Potassium | 18* | -1.45 | 0.94 | 0.22 | -1.92 to -0.98 | <0.0001 | 0.75 | 0.44 to 0.90 | 27.77 | 14.19 | -0.05 | 55.48 | 3.5 - 5.3mmol/L | No | No | No | 0 |
| Urea | 36 | -0.77 | 0.55 | 0.09 | -0.96 to 0.58 | <0.0001 | 0.96 | 0.93 to 0.98 | 14.46 | 10.15 | -5.43 | 34.35 | 2.5 - 7.8 mmol/L | No | Yes | No | 1 |
| Creatinine | 36 | -9.14 | 14.38 | 2.40 | -14.00 to 4.27 | 0.0005 | 0.83 | 0.69 to 0.91 | 11.63 | 17.89 | -23.43 | 46.70 | 60-120 umol/L | No | Yes | No | 1 |
| **Bilirubin** | 59 | -0.41 | 0.85 | 0.11 | -0.63 to 0.18 | 0.0005 | 0.98 | 0.97 to 0.99 | 5.88 | 10.38 | -14.47 | 26.23 | 0 – 20 umol/L | No | Yes | No | 1 |
| **ALP** | 59 | 4.10 | 55.92 | 7.28 | -10.47 to 18.68 | 0.2561 | 0.89 | 0.82 to 0.93 | 1.61 | 19.90 | -37.39 | 40.61 | 30 - 130 U/L | Yes | Yes | No | 2 |
| **ALT** | 59 | -0.70 | 3.66 | 0.48 | -1.65 to 0.26 | 0.7827 | 0.98 | 0.97 to 0.99 | 2.75 | 13.20 | -23.12 | 28.62 | 8 – 40 IU/L | Yes | Yes | No | 2 |
| **Total protein** | 58 | 0.26 | 2.92 | 0.38 | -0.51 to 1.03 | 0.5032 | 0.86 | 0.78 to 0.92 | -0.37 | 3.92 | -8.07 | 7.32 | 60 - 80 g/L | Yes | Yes | Yes | 3 |
| **Total IgE** | 35 | 12.23 | 34.98 | 5.91 | 0.21 to 24.24 | 0.7104 | 0.99 | 0.99 to 1.00 | -34.87 | 69.23 | -170.6 | -100.80 | 3-150 IU/mol | Yes | Yes | No | 2 |
| **Vit D** | 26* | -0.89 | 12.72 | 2.50 | -6.02 to 4.25 | 0.7258 | 0.90 | 0.78 to 0.95 | -7.26 | 53.68 | -112.5 | 97.96 | 50 - 75 nmol/L - adequate<br>>75 nmol/L - optimal | Yes | Yes | No | 2 |
| **CRP** | 37 | 1.00 | 4.89 | 0.80 | -0.63 to 2.63 | 0.2213 | 0.99 | 0.99 to 1.00 | -0.41 | 4.30 | -8.83 | 8.00 | <100mg/L | Yes | Yes | Yes | 3 |
| **Albumins** | 59 | 0.00# |  |  |  | 0.4070 | 0.75 | -0.61 to 0.85 | -0.22 | 6.15 | -12.27 | 11.84 | 35 - 50 g/L | Yes | No | No | 1 |

Values in red symbolise significant difference between the tests by P values <0.05, poor correlation ( $r$ or $r_s$ <0.8) , or wide Limits of agreement.
§ Non-parametric statistics applied for non-Gaussian distribution: Wilcoxon matched pairs signed rank test, non-parametric Spearman correlation $r_s$

To evaluate patient and clinician experience and their likelihood to promote the use of capillary blood sampling, the Net Promoter Score (NPS) was applied. The NPS presents a result from −100 (all end-users are detractors) to +100 (all end-users are promoters) by subtracting the percentage of detractors from the percentage of promoters.^16^ Users are asked a single question and rate on an 11 point scale from 10 *(Extremely likely)* to 0 *(Not at all likely)*. Results of NPS can be benchmarked against scores obtained about other products from different domains.

## RESULTS

### Phase 1: Validation study

117 of 124 patients recruited completed the study by providing both capillary and venous blood samples. Variation in number of paired samples was dependent on both numbers recruited for each group, or either haemolysis of the blood samples or insufficient blood in the sample to allow for analysis of all the tests within the subgroups.

Statistical analysis revealed that HbA1c (%), CRP and Total Protein met *a priori* criteria to demonstrate interchangeability to venous blood (Table 1). HbA1c (IFCC) had 95% confidence intervals just outside of 10% difference but a small bias (average difference between results) (~1%) suggesting high clinical usability. A number of tests such as ALT, ALP, vitamin D, Total IgE had non-significant difference by paired t-test, with strong correlation (r or r_s_ >0.8) but 95% confidence intervals greater that 10% by Bland-Altman analysis (Table 1, and Figure 3). ALT and ALP had small bias however (2.75% and 1.61% respectively) suggesting good clinical usability, with vitamin D and Total IgE showing a larger bias (7.3% and 34% respectively). Bland-Altman analysis is also shown in Supplementary Table 1 and Supplementary Figure 6. Paired t comparison is shown in Figure 2, and correlation analysis is shown in Figure 4.

**Figure 2:**
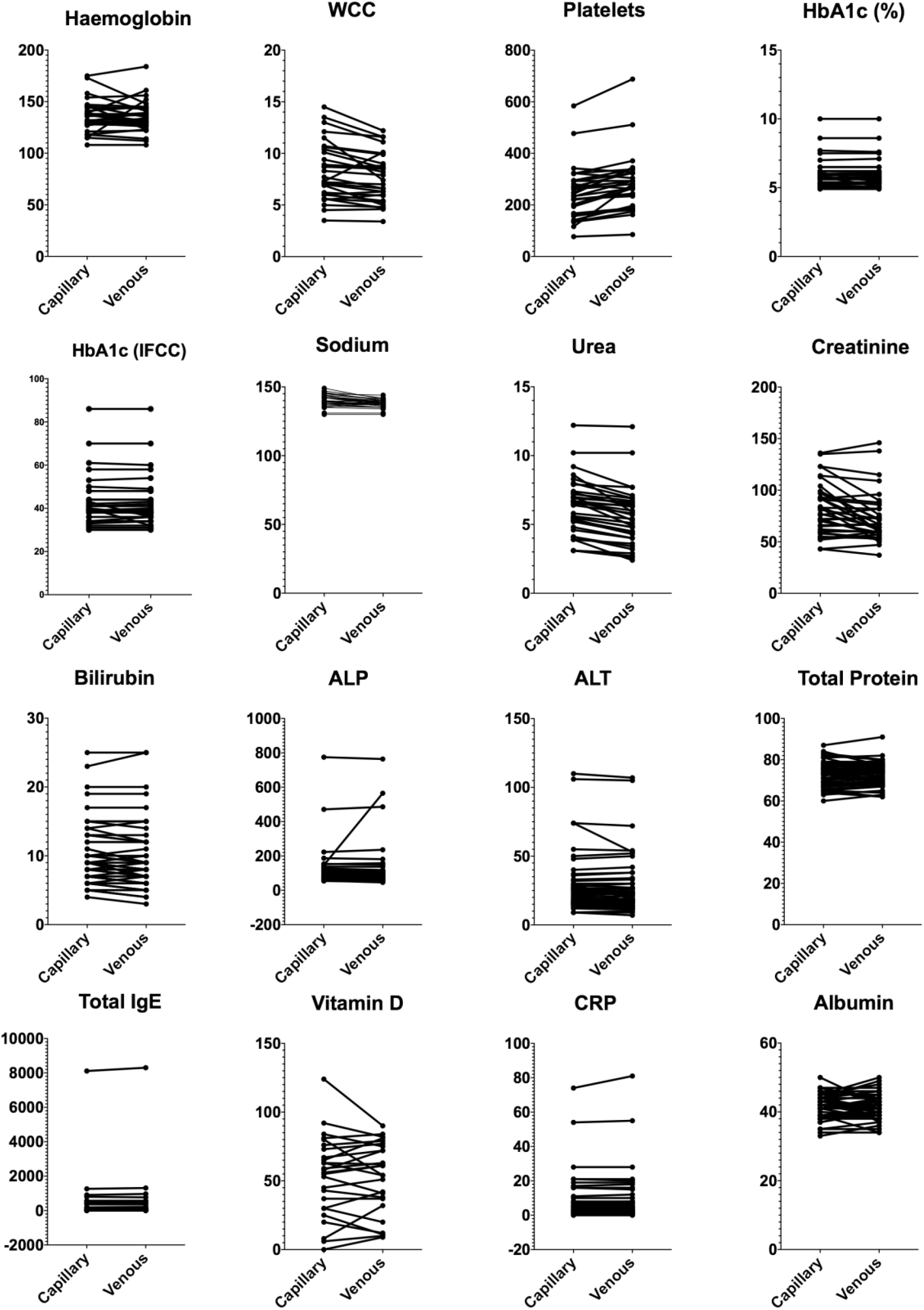
Paired T comparison for capillary and venous samples. The paired t-test enables comparisons of the means of these two methods. Key: ALP -Alkaline phosphatase, ALT -Alanine Transferase, CRP - C-Reactive protein, IgE-Immunoglobulin type E, Hb -Haemoglobin, HbA1c - glycated haemoglobin, WCC - White cell count.

**Figure 3:**
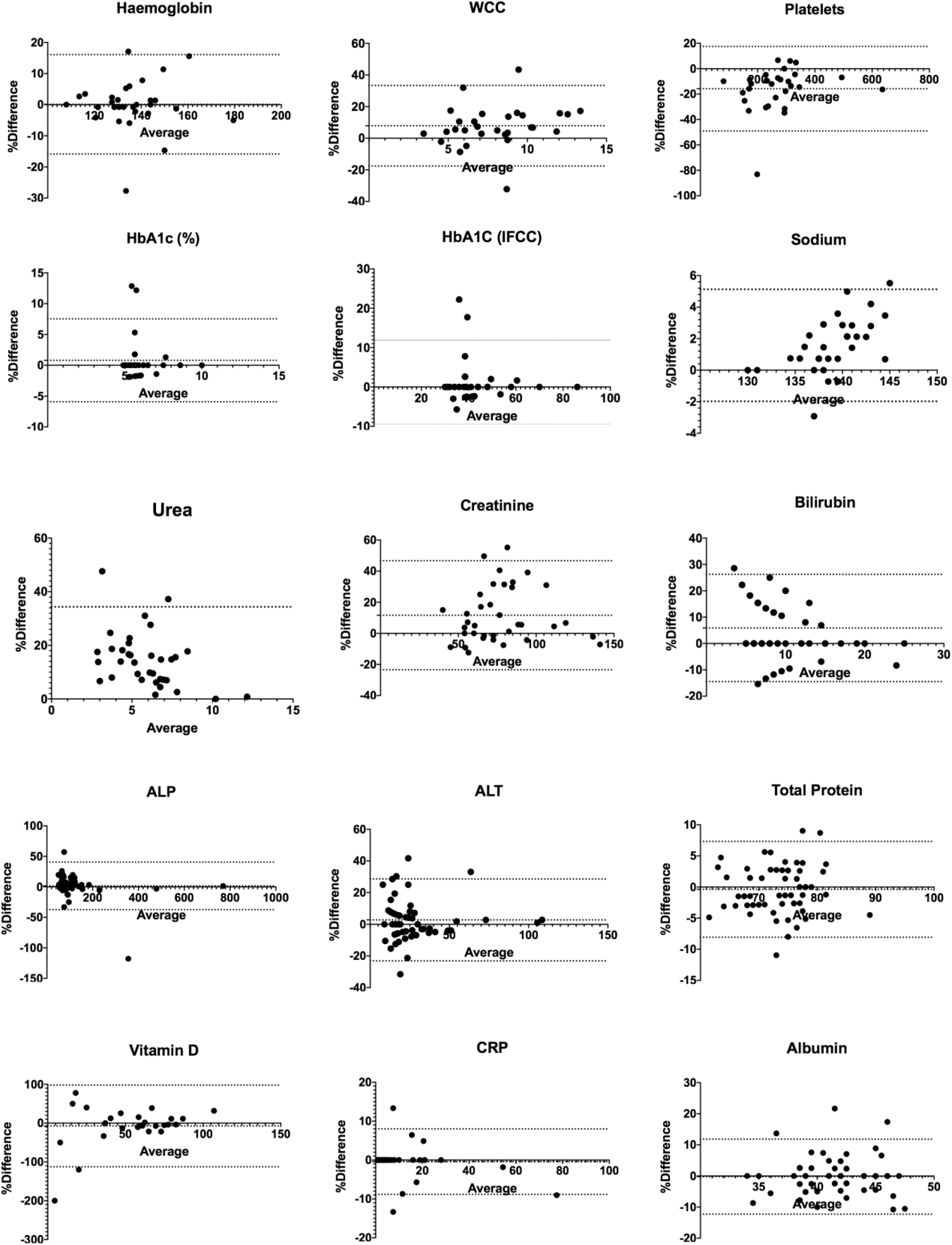
Bland Altman comparison for capillary and venous samples-% difference. The % difference between the two measurements (capillary and venous) per test is plotted against the mean of the two measurements. A 10% overall difference (either side of the zero line) between tests shows the two tests can clinically be used interchangeably. The dotted the lines represent the upper and lower limits of the 95% limits of agreement (LOA) (mean difference (bias) plus or minus 1.96 times its SD). Narrow limits of agreement show good agreement, wide LOA show poor agreement. The line of perfect agreement is the line crossing zero. The closer the points are to zero, i.e. the smaller the scatter of points from zero, the stronger the agreement between the two measurements. Key: ALP -Alkaline phosphatase, ALT -Alanine Transferase, CRP - C-Reactive protein, IgE-Immunoglobulin type E, Hb -Haemoglobin, HbA1c - glycated haemoglobin, Vit - Vitamin, WCC - White cell count.

**Figure 4:**
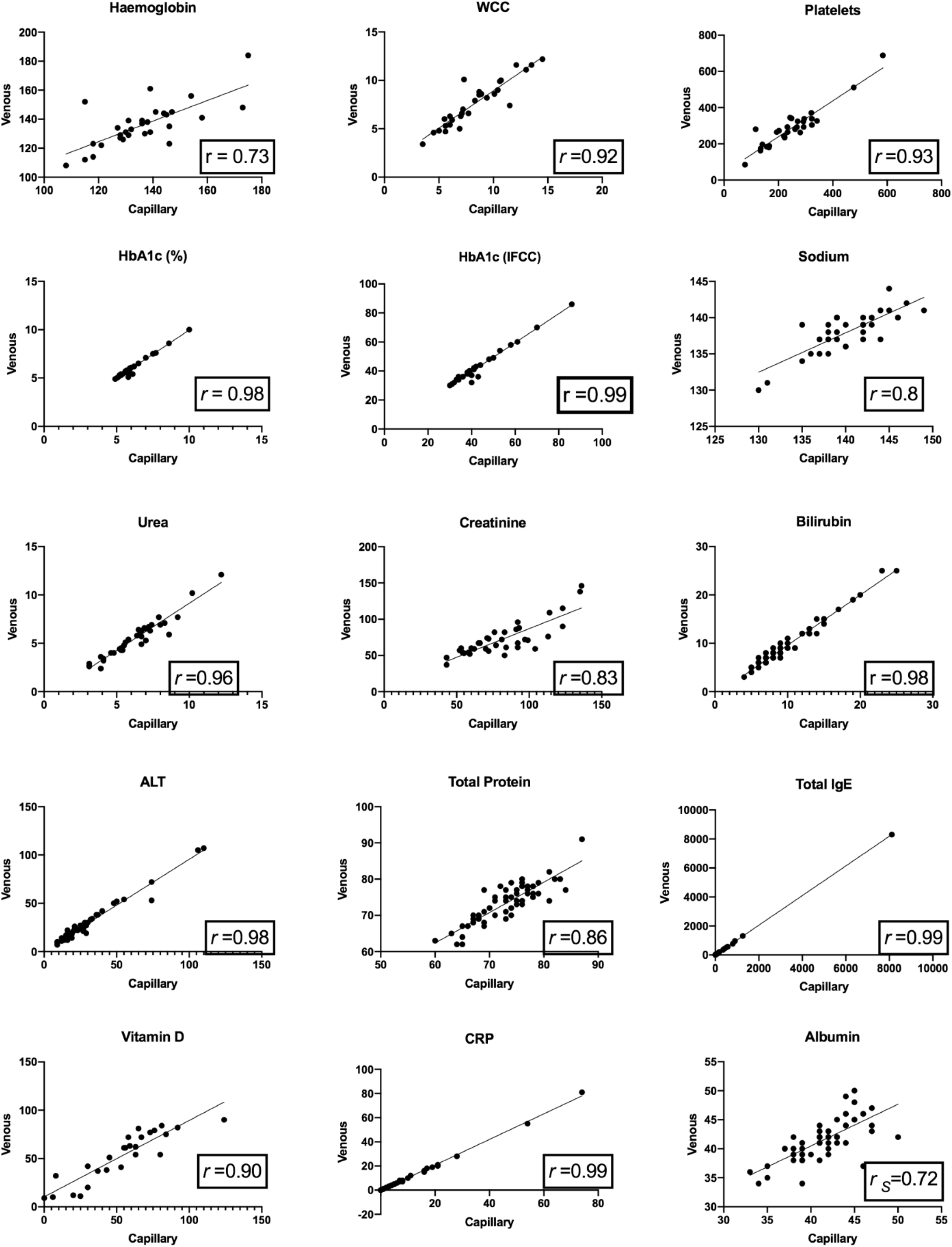
Correlation of measurements (xy analysis). The Line shows line of best fit where the simple linear regression test has been applied. *Key r*=Pearson’s correlation coefficient. R s =Spearman’s Rank correlation coefficient. ALP -Alkaline phosphatase, ALT -Alanine Transferase, CRP - C-Reactive protein, IgE-Immunoglobulin type E, Hb -Haemoglobin, HbA1c - glycated haemoglobin, Vit - Vitamin, WCC - White cell count.

Potassium showed significant difference in paired t-test, weak correlation and wide limits of agreement in Bland-Altman analysis suggesting capillary blood sampling is unreliable compared to venous blood. A number of other tests showed strong correlation, but significantly differed in paired t-test with wide limits of agreement in Bland-Altman analysis (e.g. Bilirubin, WCC, Platelets, Urea, Creatinine). This suggests although significantly different compared to venous blood, capillary testing, given the strong correlation, may potentially have a role in longitudinal monitoring but further analysis is necessary to understand clinical usability and longitudinal variability.

The majority of capillary samples went through the Royal Mail postal system via first class post and were received into the hospital laboratories for analysis within 1 day (see supplementary table 2; Transit duration (days) of capillary samples) with a median transit time of 1 day). Samples with longer transit time did not show a greater Coefficient of variation (CV) than shorter in-transit times, but it was apparent in this study that samples that were in transit for longer than 4 days had a greater propensity to clot or haemolyze (See supplementary table 2), which could render the test invalid for analysis. Coefficient of variation, however, was not greater with haemolysis index values of greater than zero.

### Phase 2: Clinical pathway mapping

Parallel to the validation study, a 4-week prospective analysis of blood testing in outpatient clinics was undertaken with a total of 13 clinics and 181 patients. 63 patients underwent blood testing (35%). Of the patients who required blood testing in clinic, 6 patients were unable to have blood tests when required; 4 due to unavailability of phlebotomy services and 2 because tests were time specific. 16% (n=10) of patients had to be contacted after clinic by the clinical team following blood results to action a change in clinical plan such as recommendation of follow-up GP visit / change in prescription etc.

Indication and predictability were assessed for each patient based on pre-existing medical history. 65% (n=41) of blood tests were predictable prior to clinic appointment. Of predictable blood tests, 78% (n=32) (and so 51% of all blood tests) were tests that were deemed interchangeable or clinically acceptable using capillary blood sampling based on the validation study.

Specific groups of patients were identified who were predicted to require blood tests during their routine clinic appointments and a novel clinical pathway was designed (see supplementary Figure 1). Once received, the patient performed the test at home. They then return-posted the capillary blood test (CBT) sample to the hospital where the hospital laboratory processed the sample. This ensured results were available in advance of clinic appointment. Over a two-week period and six outpatient clinics, 26 patients were contacted by phone to be involved in the remote capillary blood sampling pilot. Of those, 7 patients were unable to be contacted by phone and 5 declined. 14 patients agreed to take part in the pilot and were sent the *Thriva®* finger-prick capillary blood sampling device in the post. Of these, 8 patients had blood tests available to them when they attended clinic. As part of the ‘Study’ phase of the PDSA cycle (See Supplementary Figure 2), we analysed system failures and barriers to use. There was a range of reasons for the other patients’ results not being available in clinic. One patient put the label on the packaging and not directly on the blood bottle, two patients found the process difficult to collect the capillary blood, one patient received the sampling kit too late to send in time for the clinic appointment, and one patient sent the capillary blood sample but it had not arrived in time for their clinic appointment.

The NPS score showed very good usability of the capillary test (Figure 5). Feedback from study participants were obtained and 5 patients were “very satisfied” with the home blood testing kits, 2 were “quite satisfied” and 1 was “neither satisfied nor dissatisfied”. All 8 patients either “very much agreed” or “agreed” with the following statements:

1. Did you feel the process resulted in better decision making and planning?
2. Did you feel more involved in your care?
3. Did doing the tests at home save time in clinic?

**Figure 5:**
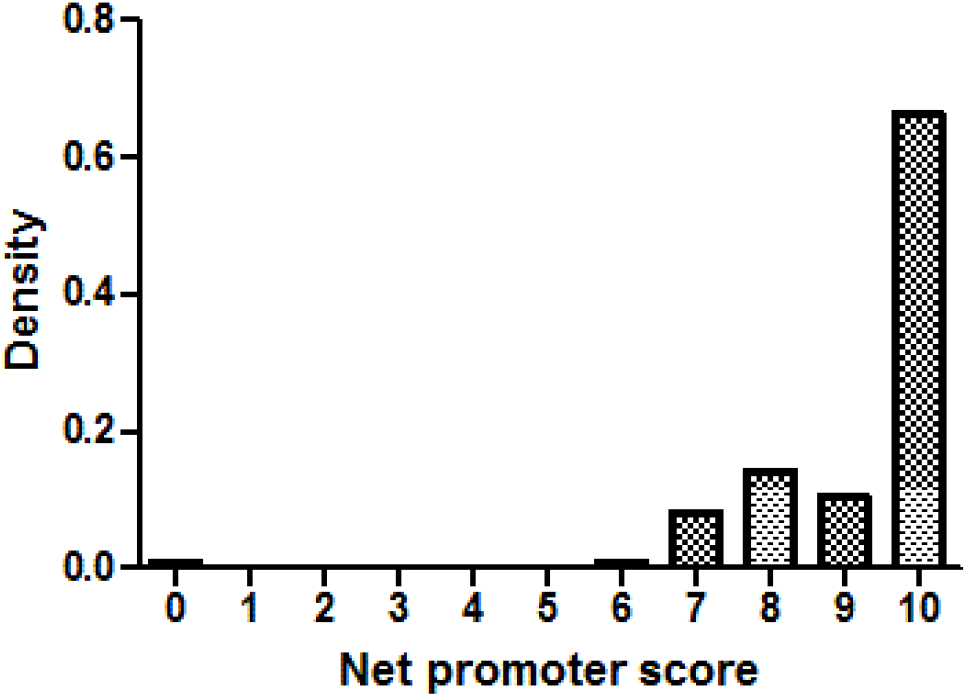
Summarises the usability results from the Net Promoter Scores-patient experience. 0 indicates the most negative experience, and 10 indicates the most positive experience.

For the 8 patients where contemporaneous blood results were available to the treating clinician and following completion of the clinic visit, they provided feedback from a clinician perspective. In all 8 clinical interactions, it was either “very much agreed” or “agreed” on the following questions: Do you feel that having the blood results available helped shared decision making and care planning? Did the process change your clinical interaction with the patient? In all but 1 clinical interaction, it was felt that the process helped save time in clinic.

## DISCUSSION

The use of remote capillary blood testing to streamline chronic disease outpatient care has not been previously evaluated. In this prospective cross-sectional observational bi-phased study, we evaluate the accuracy and usability of remote capillary blood sampling and determine the potential impact in outpatient clinical pathway mapping and potential usability to facilitate virtual cohort monitoring as required within the current COVID-19 pandemic and afterwards.

The validation study revealed interchangeability between venous and capillary blood testing in a number of blood tests including HbA1c, total protein and CRP testing. Further capillary tests such as ALT, ALP, Total IgE (Immunoglobulin E), and Vit D (Vitamin) appear to have a place for use in clinical practice for remote monitoring given strong correlation, however the higher levels of variation with Total IgE and vitamin D in particular suggests further confirmatory analysis will be required. Importantly, this study confirms that capillary results for certain tests such as potassium appear unreliable and should not be used within clinical practice, as delays in analysis affects the final reading.^17-20^ Interchangeability score was low for creatinine testing, but the Jaffe method used for creatinine analysis at our labs could have some influence in this^21^ as it is more susceptible to interferences compared to enzymatic assays^22^. Capillary and venous creatinine blood comparisons may produce differing results where this study is repeated with creatine sampling done in labs with more specific enzymatic assays.

A number of tests (e.g. Bilirubin, WCC, Platelets, Urea, Creatinine) appear significantly different from venous blood but highly correlated suggesting a possible role in intra-patient longitudinal monitoring in the outpatient setting for patients with long term clinical diseases. Differences in capillary and venous Haemoglobin counts is long established but the mechanism for this is unknown^23^. WCC and platelets have been similarly affected, with some studies describing non-significant differences^24^ and some recognising that there are differences^25^ with greater precision achievable with venous blood^23^. As in our study, good correlation has been observed in the literature with capillary and venous comparisons of WCC albeit in different patient populations such as oncology^24^ or healthy athletes^26^. Though it cannot be said that they are interchangeable, clinical utility can be considered for a number of tests (e.g. Sodium and Bilirubin) as good correlation and low bias was demonstrated. Similarly, albumin, and Haemoglobin had small bias and Bland-Altman in the former was close to the 10% difference. Results should however be interpreted with caution and further analysis with larger numbers is required to understand its clinical utility where interchangeability is desired.

Outliers exist in the data set that could possibly skew the data and potentially skew the agreements and associations between venous and capillary samples. This is evident in results for Haemoglobin and Alkaline Phosphatase for example (see Figure 3). These were not excluded from the analysis because at this stage the cause(s) of the outlier(s) are unknown, and could be technical or human factor related. Further work on human factor and process analysis will be required to understand cause and potential impact. This will be key to enable adoption within novel clinical pathways, given the absence of a contemporaneous venous result for comparison.

As a pilot study, the results are limited by size and its cross-sectional nature. In addition, any choice of statistical analysis of agreement has limitations. Although using *‘a-priori’* percentage difference limits within Bland-Altman analysis can be useful as a measure of agreement for tests where the normal range is wide (e.g. Total IgE, which can range from single figures to thousands), it can be problematic in dealing with clinically insignificant change within normal range albeit with a high ‘percentage’ change. In translating to a clinical setting, larger multicentre studies will be required to analyse ‘clinically meaningful change’ and to determine use within screening protocols(e.g. vitamin D, Haemoglobin, etc), analysing sensitivity and specificity. Within chronic disease, as the majority of testing relates to longitudinal monitoring, further longitudinal analysis is required, but the high correlation coefficients seen in a number of tests (e.g. liver function, CRP, Total IgE, HbA1c) suggest capillary testing may be an extremely useful practical tool for clinicians. These tests are highly clinically relevant and have a range of uses, such as disease monitoring and drug toxicity monitoring. Within chronic respiratory disease care, given a large proportion of ongoing monitoring relates to allergy, infection and hepatotoxicity this is particularly useful. The ability to accurately remotely monitor HbA1c is potentially also helpful across chronic disease, but also in tertiary respiratory care where ~50% of adults with Cystic Fibrosis have diabetes.^27^

Our study shows good usability of remote capillary testing. These results are promising for real life implications as the capillary samples were all posted using the UK Royal Mail postal service to reflect a real clinical pathway. Results showed the tests were largely stable within the postal system for up to 6 days, and could be used provided the samples have not haemolysed. The recommendation if haemolysis occurs will be to request a repeat capillary sample.

There is a recognised inefficiency in the form of failure demand in healthcare when clinical results need to be reviewed and plans amended after the time of clinical interaction. Our analysis of clinic blood testing showed that 16% of patients who underwent blood testing needed to be contacted at a later date by a healthcare professional to amend the initial clinical plan put in place at the time of the clinic appointment. This is time-consuming and costly for both healthcare professionals and patients. Furthermore, we revealed that over half of the blood tests performed in our chronic respiratory disease clinics could potentially both be predicted and provided by remote capillary blood testing. This is particularly pertinent in the current COVID-19 pandemic, where a large percentage of testing could be predicted and reliably achieved remotely to facilitate virtual consultation in a shielded population.

To show proof-of-principle we designed a small pilot study to target inefficiency and streamline blood monitoring in chronic respiratory disease care. This showed good subjective evidence that patients felt more involved in care and healthcare professionals found it saved time in clinic. Within our pilot study, we rolled out an early model to learn from the successes and barriers through a PDSA (Plan Do Study Act) cycle^28^ of learning (Supplementary Figure 2) and map out the follow on actions to be undertaken (‘’Act’’ Phase). This work is ongoing to enable a streamlined integration within clinical practice. Further evaluation is required to determine impact on clinical pathways within a complete virtual remote care platform, alongside health-economic modelling to assess impact upon time, cost and clinical decision-making outcomes.

Though the initial focus of this work was within a face-to-face outpatient clinic setting, we recognise the implications in the current COVID-19 pandemic to enable rapid expansion of remote chronic disease care for highly susceptible cohorts. Remote capillary blood test monitoring is a very attractive solution to maintaining continuity of healthcare provision whilst improving patient experience by reducing socioeconomic impact, mitigating infection risk in highly vulnerable individuals and enabling contemporaneous clinical information at time of review. We have developed a proposed novel clinical pathway (see Supplementary Figure 1) to enable use within either an outpatient, ambulatory or completely virtual telemedicine scenario for further larger-scale validation.

In summary, in this pilot study, we analyse the accuracy of remote capillary blood tests compared to standard venesection and show usability within novel clinical care pathways to facilitate remote chronic disease monitoring with high applicability to the current COVID-19 pandemic and beyond.

## Data Availability

All data referred to in the manuscript is available. Correspond with corresponding author.

## Patient consent for publication

Non required

## Ethics approval

North West - Haydock Research Ethics Committee (REC reference: 18/NW/0491)

## Table of abbreviations

| *Abbreviation* | *Explanation* |
| --- | --- |
| *ABPA* | Acute Bronchopulmonary Aspergillosis |
| *CF* | Cystic Fibrosis |
| *A. Fumigatus* | *Aspergillus fumigatus* |
| *COPD* | Chronic obstructive pulmonary disease |
| *ILD* | Interstitial Lung Disease |
| *Pulm HTN* | Pulmonary hypertension |
| *ALP* | Alkaline phosphatase |
| *ALT* | Alanine Transferase |
| *CRP* | C-Reactive protein |
| *IgE* | Immunoglobulin type E |
| *Hb* | Haemoglobin |
| *HbA1c* | Glycated haemoglobin |
| *Vit* | Vitamin |

